# Postprandial glycaemic response to white and wholemeal bread consumption between normal weight and overweight/obese healthy adults

**DOI:** 10.1101/2025.01.04.25319987

**Authors:** Honglin Dong, Abbie Colosimo, Yizhi Xu

## Abstract

Obesity and the increased postprandial glycaemic response (PPGR) are risk factors for type 2 diabetes. Few studies have explored the association of body weight with PPGRs. The study aimed to investigate the PPGR between healthy adults with normal weight and overweight/obesity to two commercially available breads (white and wholemeal) with different dietary fibre contents. In this acute randomised crossover trial, 20 healthy adults (10 normal weight, 10 overweight/obese) consumed two slices of white (100 g, fibre 3.6 g) or wholemeal bread (88 g, fibre 5.6 g) alongside 150 ml of orange juice and 10 g butter on separate visits in random order after fasting for 8-12 hours. The blood glucose concentration was measured fasted, 30 min, 60 min, 90 min and 120 min postprandially by finger pricks. Information on age, gender, ethnicity, body mass index (BMI), and body fat percentage were collected. Two-way repeated measures ANCOVA was used for controlling for age, gender, ethnicity and body fat percentage, and results showed no significant difference was observed in fasting blood glucose concentrations (F(1, 14)=2.968, P=0.107), incremental areas under the curve (iAUCs) (F (1, 14) = 0.702, P=0.416) and peak values (PVs) (F (1, 14) = 0.507, P=0.488) between participants with normal weight and overweight/obesity, or in fasting blood glucose concentrations (F(1, 14)=0.007, P=0.964), iAUCs (F (1, 14) = 0.008, P=0.929) and PVs (F (1, 14) = 0.036, P=0.851) between white and wholemeal bread consumption. The BMI or body fat percentage was not associated with iAUCs or PVs regardless of bread type adjusted by age, gender and ethnicity. Our results did not find that body weight is associated with PPGR in healthy adults, and the wholemeal bread used in the current study did not deliver the health benefit of attenuating PPGR compared with white bread consumption despite the higher dietary fibre content.

## Introduction

Postprandial glycaemic response (PPGR) is an independent risk factor for type 2 diabetes mellitus (T2DM) (Abdul-Ghani et al., 2008) and a loss of postprandial glucose control is the first metabolic alteration detectable in the progression of prediabetes to T2DM (Monnier, Lapinski & Colette, 2003). It is well-known that T2DM is a multifactorial disease, and overweight and obesity are major risk factors for T2DM (Klein et al., 2022). Excessive fat content in adipose tissue will cause lipolysis and result in the release of free fatty acids to the circulation which promotes muscle and hepatic insulin resistance as well as impaired insulin secretion of the ß-cell in the pancreas (DeFronzo et al., 2015; Shulman et al., 2014). However, previous evidence shows inconsistent results regarding the association between body weight and PPGR. An acute crossover human trial in healthy participants (Perälä et al., 2011) found there was no significant difference in the incremental area under the curve (iAUC) of PPGR up to 2 hours between normal weight and overweight/obese participants regardless of low or high glycaemic index meal consumption. Another study with a similar study design found that adults with central obesity (determined as a waist circumference ≥102 cm or body mass index, BMI ≥30 kg/m^2^) had significantly higher PPGR presented as AUC than lean adults particularly between 0-60 min after standardised breakfast consumption with either preload of a whey protein or control drink (Smith et al., 2021).

Bread is a staple food in the UK, among which white bread is the most consumed bread in the UK followed by wholemeal or wholewheat bread (Wunsch, 2022), although wholemeal bread is regarded as healthier than white bread and recommended to the public over white bread due to its higher dietary fibre content and nutrients such as phytochemicals and minerals including magnesium, selenium and copper (Lockyer & Spiro, 2020). However, few studies have investigated the relationship between body weight and PPGR after consuming these two common food items, white and wholemeal bread. In addition, we were also very interested in whether in real life wholemeal bread does reduce the PPGR compared with white bread consumption as expected.

The primary outcome of the study was to explore whether overweight and obese people had a higher PPGR compared with people with normal weight after consuming commercially available white and wholemeal breads in UK supermarkets. The secondary outcome was to investigate whether the consumption of wholemeal bread could attenuate PPGR compared with white bread consumption regardless of body weight.

## Methods

This acute randomised crossover trial was approved by the Coventry University Ethics Committee (Ref P136390) and conducted between May and July 2022 at the School of Life Sciences at Coventry University, the United Kingdom. All participants provided written consent before participating.

### Participants

Twenty participants were recruited using purposive and convenience sampling via circulating recruitment advert via email and word of mouth among the university staff and students. Normal weight (BMI 18.5-24.9 kg/m^2^) and overweight/obesity (BMI ≥ 25 kg/m^2^) were defined according to NICE (2023).

The sample size (n=20) was referred to a similar study design to detect the significant di erence in PPGR among different breads with a level of α 0.05 and β 0.08 (Zafar et al., 2020). The inclusion criteria were healthy adults of 18-50 years. The exclusion criteria were participants with diabetes, digestive system diseases, BMI < 18.5 kg/m^2^, coeliac disease, other chronic diseases, blood clotting issues, and those who could not consume the study meals due to food allergy or other reasons. The eligibility of the participants was screened via a health and lifestyle questionnaire.

### Study design

Eligible participants had two visits (at least 48 hours of interval). On each visit, participants were asked to fast for 8-12 hours starting from the night before their visit (only water is allowed). Participants were provided meals with different types of bread, white bread (WB) and wholemeal bread (WMB) in random order (participants drew lots to decide on bread consumption sequence on their first visit). Participants consumed two slices of white or wholemeal bread alongside 150 ml of pure orange juice and 10 g butter. All food and drink items were purchased from Tesco supermarket (Welwyn Garden City, United Kingdom). The details of the meal composition and nutrient information are shown in **Table 1**.

**Table 1.** Details of meal components and nutrient contents of white and wholemeal bread meals.

|  | White bread meal | Wholemeal bread meal |
| --- | --- | --- |
| Bread (2 slices) (g) | 100 | 88 |
| Pure orange juice (ml) | 150 | 150 |
| Butter (g) | 10 | 10 |
| Total energy (kcal) | 343.4 | 328.2 |
| Carbohydrate (sugar) (g) | 60 (17.8) | 48 (17.4) |
| Total dietary fibre (g) | 3.6 | 5.6 |
| Protein (g) | 9.2 | 11.1 |
| Fat (g) | 9.4 | 11.8 |

The breads and the orange juice were produces of TESCO (Welwyn Garden City, United Kingdom) and butter was produced by Arla Foods Ltd. (Leeds, UK). All food and drink items were purchased from local supermarkets. The nutrition content was obtained from the provided nutrition information on the package.

Postprandial blood glucose was measured at 0 (fasting), and 30, 60, 90 and 120 min by finger prick performed by the researcher using Biosen Blood Glucose/Lactate Analyser (EKF Diagnostics, Cardiff). Participants were asked to consume the meal within 10 min, keep sedentary, and not eat or drink during the study period. Mobile alarms were used to ensure blood samples were collected on time. The body height was measured by a stadiometer, and the body weight and fat percentage were measured by Tanita MC-980MA PLUS (Tanita Company, Tokyo) before meal on the first visit only. In addition, the participants were asked to avoid intensive physical activity and alcohol and have a good night’s sleep the night before their visit day. The names, email addresses and mobile numbers were collected for appointment purposes and the self-reported demographic variables (age, gender and ethnicity) were collected and considered confounding factors.

The researcher who conducted the study and the participants were aware of what bread was consumed for each visit (non-blinded).

### Data analysis

Categorical data including gender (male and female), ethnicity (white Caucasian and non-white Caucasian) and body weight categories (normal weight and overweight/obese) were presented as frequency (n) and percentage (%). Continuous data including age, BMI, body fat percentage, blood glucose concentration etc. were presented as mean ± standard deviation (SD). The postprandial glycaemic response was reported as iAUC and the peak value (PV) of the blood glucose concentration after the meals for up to 2 hours. The iAUC was calculated using approximated trapezoidal numerical integration (Chlup et al., 2008), and only the incremental area above the fasting level was included.

The iAUCs, PVs and the fasting blood glucose concentrations between body weight categories (normal weight and overweight/obese) and between the two bread consumptions (white bread and wholemeal bread) were analysed by two-way repeated-measures Analysis of Covariance (ANCOVA), bread as a within-subjects variable, body weight categories as a between-subjects variable, and age, gender, ethnicity, body fat percentage as covariates. Partial correlation was used to analyse the association of BMI and body fat percentage with iAUCs and PVs after white and wholemeal bread consumption separately by considering age, gender and ethnicity as confounding factors. Normality of the continuous data (fasting blood glucose concentration, iAUC, PV, BMI, age, body fat percentage) were carried out using Kolmogorov-Smirnov test. All continuous variables were normally distributed. The statistical significance was set up at P ≤ 0.05 two-tailed.

## Results

### Basic information about the participants

Figure 1 shows the participant flow chart. Twenty-two participants were screened among which one had a BMI < 18.5 kg/m^2^, another had T2DM. Therefore, 20 participants were eligible and recruited, and all completed the study.

**Figure 1.**
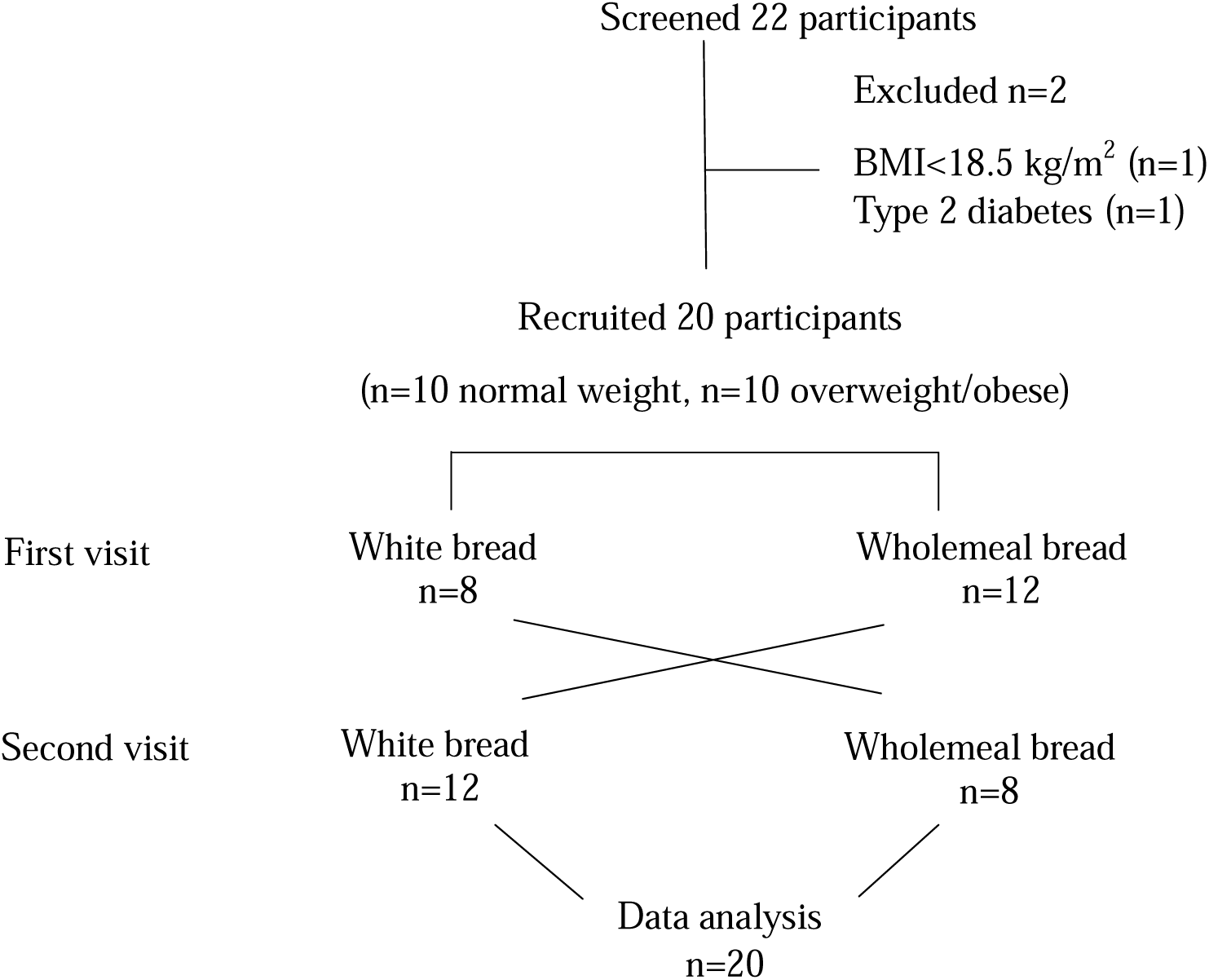
Participant flow diagram.

**Table 2** shows the characteristics of the participants. There were 10 normal weight (BMI 21.7 ± 2.8 kg/m^2^, body fat percentage 21.0 ± 4.3 %) and 10 overweight/obese participants (BMI 28.9 ± 3.5 kg/m^2^, body fat percentage 30.1 ± 5.7 %), among whom 12 were females and 8 were males. Thirteen participants were white Caucasians and 7 were non-white including 3 Chinese, 2 Indians, and 2 black Africans. The mean age was 32.7 ± 10.2 y (18-48 years); the mean BMI was 25.3 ± 4.9 kg/m^2^ (18.5-31.2 kg/m^2^), and the mean body fat percentage was 25.6 ± 6.8 % (from 12.5 % - 40.5 %).

**Table 2.** Participants’ characteristics.

| Participants | n (%) | Age (year) |  | BMI (kg/m<sup>2</sup>) |  | Body fat (%) |  |
| --- | --- | --- | --- | --- | --- | --- | --- |
|  |  | Mean | SD | Mean | SD | Mean | SD |
| Total | 20 (100%) | 32.7 | 10.2 | 25.3 | 4.9 | 25.6 | 6.8 |
| Female | 12 (60%) | 32.3 | 9.9 | 25.0 | 5.6 | 27.8 | 6.6 |
| Male | 8 (40%) | 33.1 | 11.1 | 25.7 | 3.8 | 22.3 | 5.9 |
| White | 13 (65%) | 32.2 | 10.0 | 25.5 | 5.5 | 26.3 | 7.5 |
| Non-white | 7 (35%) | 33.6 | 11.2 | 25.0 | 3.7 | 24.2 | 5.3 |
| Normal weight | 10 (50%) | 30.3 | 9.8 | 21.7 | 2.8 | 21.0 | 4.3 |
| Overweight/obese | 10 (50%) | 35.0 | 10.4 | 28.9 | 3.5 | 30.1 | 5.7 |

BMI, body mass index; n, number; SD, standard deviation.

### Fasting glucose concentrations

The mean fasting glucose concentration was 3.2 ± 1.0 mmol/L and 3.4 ± 1.0 mmol/L before consumption of white bread and wholemeal bread meals respectively. There was no significant difference in the fasting glucose concentrations between the body weight categories (between-subject effect, F(1, 14)=2.968, P=0.107) nor between the two breads consumed (within-subject effect, F(1, 14)=0.007, P=0.964) after controlling for age, gender, ethnicity and body fat percentage (**Table 3**).

**Table 3.** Fasting glucose concentrations (mmol/L)

| Group | White bread |  | Wholemeal bread |  | P value |
| --- | --- | --- | --- | --- | --- |
|  | Mean | SD | Mean | SD |  |
| All participants (n=20) | 3.2 | 1.0 | 3.4 | 1.0 | Within-subject effect<br>(bread) 0.964 |
| Normal weight (n=10) | 3.1 | 1.1 | 3.4 | 1.1 | Between-subject effect<br>(body weight) 0.107 |
| Overweight/obese (n=10) | 3.3 | 1.1 | 3.4 | 1.1 |  |

SD, standard deviation. Two-way repeated measures ANCOVA with bread as the within-subjects variable and body weight as the between-subjects factors adjusted by age, gender, ethnicity and body fat percentage as covariates.

### PPGRs between participants with normal weight and overweight/obesity and between two bread consumptions

Figure 2 shows the PPGRs over the 2 hours between normal weight and overweight/obese groups after white (Figure 2 A) and wholemeal bread consumption (Figure 2 B). There was no significant difference in iAUCs and PVs between the two body weight groups after controlling for age, gender, ethnicity and body fat percentage (F (1, 14) = 0.702, P=0.416, and F (1, 14) = 0.507, P=0.488 respectively) (Table 4). There were no bread and body weight interactions for iAUCs and PVs.

**Figure 2.**
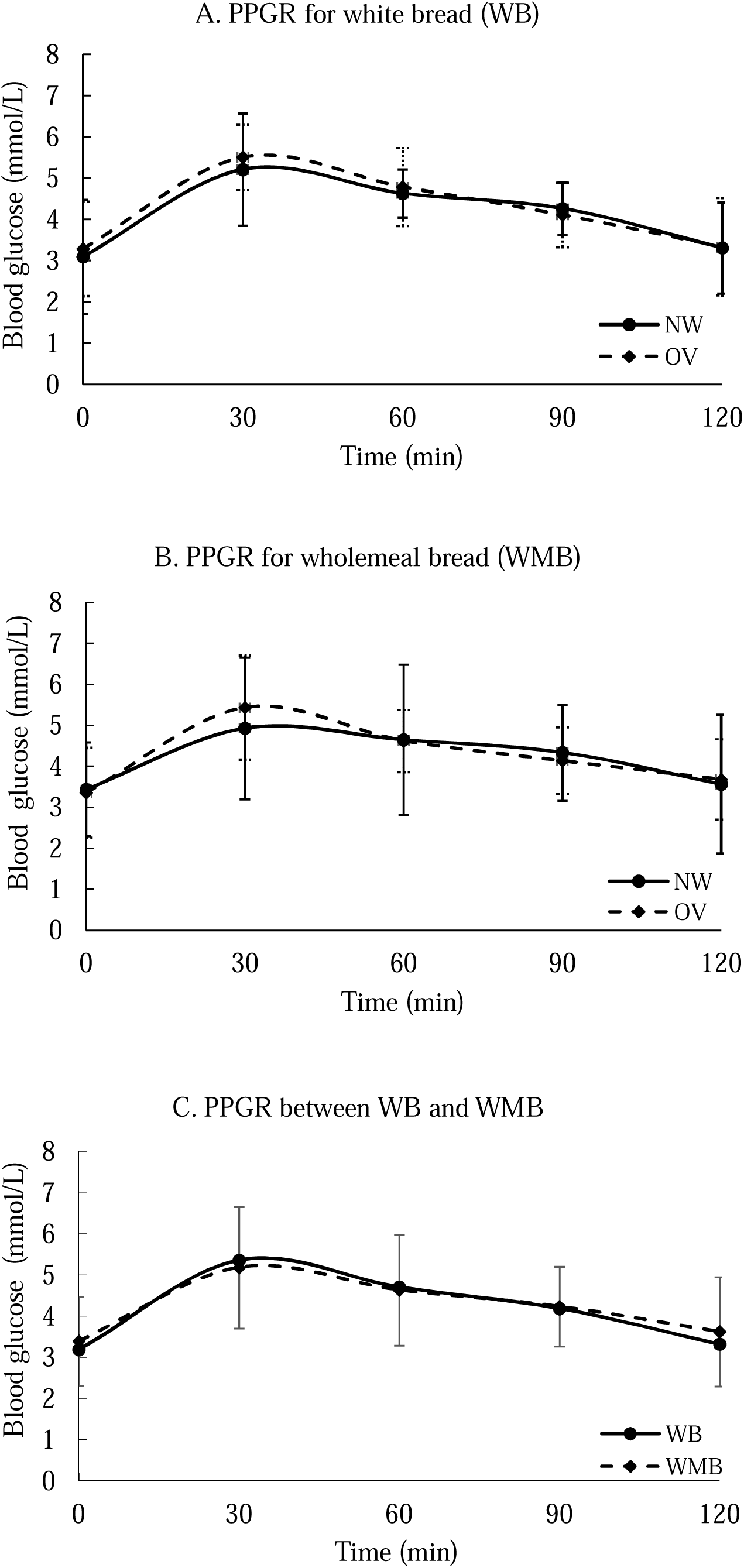
Postprandial glycaemic response. A. after white bread consumption between normal weight (NW) and overweight/obese (OW); B. after wholemeal bread consumption between normal weight (NW) and overweight/obese (OW); C. after white bread and wholemeal bread consumptions including all participants.

**Table 4.** Comparisons of iAUCs and PVs between participants with normal weight and overweight/obesity.

| Groups | Normal weight<br>n=10 |  | Overweight/obese<br>n=10 |  | P value |
| --- | --- | --- | --- | --- | --- |
|  | Mean | SD | Mean | SD |  |
| iAUC-WB (mmol □ min/L) | 147.7 | 92.0 | 137.0 | 103.3 | Body weight: 0.416<br>Bread: 0.929 |
| iAUC-WMB<br>(mmol □ min/L) | 109.4 | 121.6 | 128.5 | 60.7 | Bread*Body weight: 0.502 |
| PV-WB (mmol/L) | 5.5 | 1.0 | 5.8 | 0.8 | Body weight: 0.488<br>Bread: 0.851 |
| PV-WMB (mmol/L) | 5.3 | 1.6 | 5.4 | 1.3 | Bread*Body weight: 0.549 |

iAUC-WB, incremental area under the curve-white bread; iAUC-WMB, incremental area under the curve-wholemeal bread; PV-WB, peak value-white bread; PV-WMB, peak value-wholemeal bread; SD, standard deviation. Two-way repeated measures ANCOVA adjusted by age, gender, ethnicity, body fat percentage as covariates. The asterisk \* means the interaction between factors.

The PPGRs in all participants after consuming white and wholemeal bread was shown in Figure 2 C. There was no significant difference in iAUCs and PVs between white and wholemeal bread consumptions (F (1, 14) = 0.008, P=0.929 and F (1, 14) = 0.036, P=0.851 respectively) (**Table 4**). There were no bread and body weight interactions for iAUCs and PVs.

### Association of BMI and body fat percentage with iAUCs and PVs

There were no significant associations of BMI or body fat percentage with iAUCs and PVs regardless of white or wholemeal bread consumption after adjusting age, gender and ethnicity (**Table 5**).

**Table 5.**
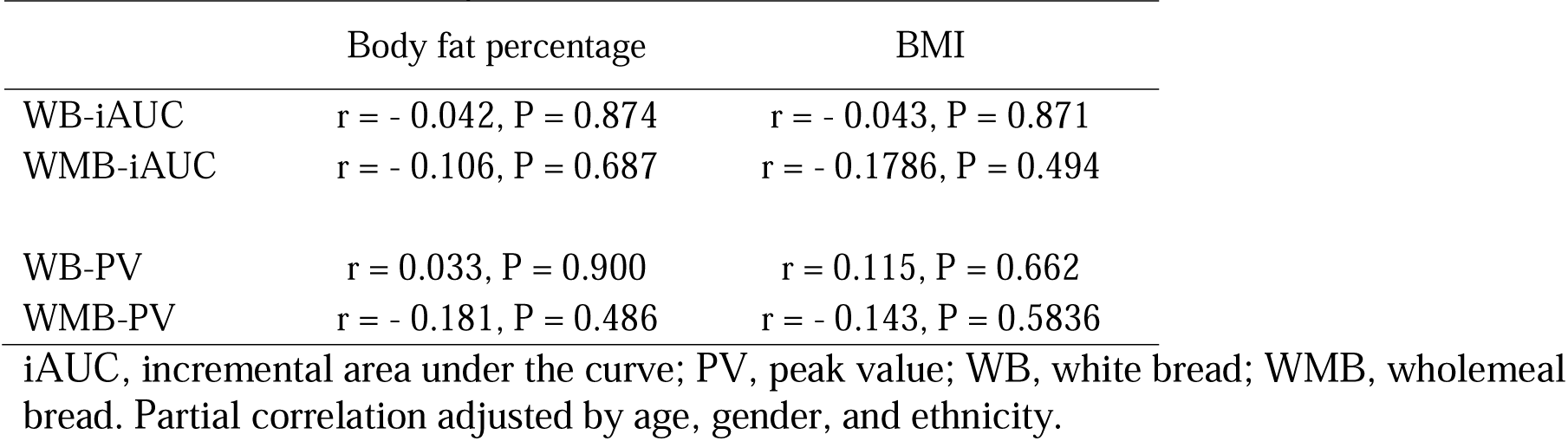
Association of body fat composition or BMI with iAUCs and PVs.

|  | Body fat percentage | BMI |
| --- | --- | --- |
| WB-iAUC | $r = -0.042, P = 0.874$ | $r = -0.043, P = 0.871$ |
| WMB-iAUC | $r = -0.106, P = 0.687$ | $r = -0.1786, P = 0.494$ |
| WB-PV | $r = 0.033, P = 0.900$ | $r = 0.115, P = 0.662$ |
| WMB-PV | $r = -0.181, P = 0.486$ | $r = -0.143, P = 0.5836$ |

iAUC, incremental area under the curve; PV, peak value; WB, white bread; WMB, wholemeal bread. Partial correlation adjusted by age, gender, and ethnicity.

## Discussion

Our result did not find a significant difference in the fasting glucose concentrations or PPGRs to the two commercially available bread consumptions, white and wholemeal (two slices each), between normal weight and overweight/obese participants. Our results were supported by several studies conducted in participants with normal glucose tolerance or with impaired glucose tolerance after consuming high- and low-glycaemic index meals (Perälä et al., 2011), in healthy lean and obese participants after consuming an oral glucose load (van Vliet et al., 2020), and in healthy young adults after consuming traditional and modified vegetarian meals (Raczkowska et al., 2019). However, some other studies showed different results. In a study investigating the postprandial effect of fresh and processed orange juice consumption on the glucose metabolism in lean and obese participants, it found that lean participants but not obese participants showed reduced iAUC (0-300 min) after fresh or processed orange juice ingestion compared with control (isocaloric sugar/acid-matched control orange-flavored drink), while obese participants secreted more insulin (> 75%) to deal with the same amount of sugar than did lean participants (Paiva et al., 2019). Another study (Leohr &Kjellsson, 2022) found that the fasting glucose and insulin concentrations in obese participants were significantly higher than in participants with normal weight, while only postprandial glucose response (not insulin response) over 13 hours following a high-fat meal was higher in obese than lean participants. However, this study lacks postprandial data between 0-4h. Moreover, a study in India found that there was a significant positive correlation between BMI and fasting blood glucose in 150 healthy adults (r=0.751, P <0.001) (Agrawall et al., 2017). The inconsistent results might be due to different meals or drinks consumed that contain various nutrient compositions and sugar contents, and different populations.

It is worth noting that fasting and postprandial insulin secretions seem better measures than fasting and postprandial glucose tests to identify early dysfunctional glycaemic metabolism in obese people. In the study by van Vliet et al. (2020), the 2-h plasma insulin AUC after glucose ingestion was significantly greater in the obese than the lean healthy adults, though there was no significant difference in 2-h plasma glucose AUC between obese and the lean groups (*P* = 0.09). In a study investigating the predictors of postprandial glycaemia, insulinaemia and insulin resistance in a pooled 108 healthy adolescents, the results show BMI was one of the significant predictors for insulin resistance but could not explain the variance in blood glucose AUC (Williams et al., 2021). Similar results were shown on childhood obesity that the correlation of glucose-based parameters (oral glucose tolerance test, fasting glucose and glycated haemoglobin or HbA1c) with BMI is poor, while the insulin secretion is significantly higher in children with obesity than in normal-weight children (Vinoy et al., 2023). This is also echoed by a debate article (Johnson, 2021) that illustrated increased insulin levels in both the fasting and fed states are the most prevalent feature of pancreas beta cell dysfunction found in obesity before insulin resistance, and well before relative glucose-stimulated insulin secretion is reduced when impaired glucose tolerance occurs. This may explain why overweight/obese healthy participants did not have an increased fasting blood glucose concentration and postprandial glycaemic response compared with participants with normal weight observed in the current study and several other studies mentioned above.

The current study compared PPGR between two breads consumed, two slices of white bread and wholemeal bread respectively that are commercially available in the UK supermarket and found no difference in PPGR represented by iAUCs and PVs was found. Interestingly, apart from the higher dietary fibre content in the wholemeal bread, the total of carbohydrates available in the wholemeal bread is 12 g less than the white bread (88 g vs 100 g carbohydrates), which is supposed to be advantageous for wholemeal bread in reducing its PPGR, however, it was not the case in our study, indicating a weaker fibre story than commonly perceived. Previous evidence indicated that increased fat or protein contents lead to a reduced but prolonged PPGR (Moghaddam et al., 2006), but the fat and protein contents in the two test bread meals of the current study were very similar, which made their influence neglectable. Belobrajdic et al. (2019) found similar results that no difference in PPGR between wholemeal and refined bread consumptions (each bread weighed 121 g) was found despite wholemeal bread having more than double the fibre content than the refined bread in 20 healthy participants. In a randomised crossover study (Hannah, Mallard & Venn, 2014), 120 healthy young adults consumed commercially available white bread and whole grain bread sandwiches that had low (49) and high (75) glycaemic indexes respectively due to different dietary fibre contents but similar available carbohydrate contents per sandwich (26 g and 27 g respectively). The results show no significant difference in the glycaemic responses (0-120 min) between white and whole grain sandwiches, represented by iAUC. A systematic review (Musa-Veloso et al., 2018) including twenty randomised controlled trials found that the consumption of wholemeal wheat including wholegrain bread and pasta, compared with white wheat, was not associated with a significant reduction in blood glucose AUC (−6.7 mmol/L⋅min; 95% CI: −25.1, 11.7 mmol/L⋅min; *P* = 0.477). Wholemeal mainly contains insoluble fibre (around 86%) (whole wheat contains a total fibre of 11.6–17.0g/100g, among which 10.2 - 14.7g are insoluble fibre, while only 1.4 - 2.3g is soluble fibre (De Santis et al., 2018). Evidence has shown that soluble fibre, particularly viscous soluble fibre, can increase the chyme viscosity to inhibit glucose absorption and reduce the gastric emptying rate, hence significantly attenuating acute postprandial glucose response (Giuntini et al., 2022), while insoluble fibre shows no effect on PPGR. For example, a study in 19 healthy postmenopausal women (Juntunen et al., 2003) found there was no significant difference in PPGR within 2 hours to three different rye breads with 50 g available carbohydrate but different fibre contents, 6.1 g for endosperm rye bread, 15.2 g for traditional rye bread, and 29.0 g for high-fibre rye bread. The difference in the fibre contents of the three breads was mainly due to different insoluble fibre contents (3.1 g, 10.9 g and 24 g respectively), while the soluble fibre contents were very similar between the breads (3.0g, 4.3g and 4.8g respectively), indicating that the insoluble components of cereal fibres are ineffective in the regulation of postprandial glycemia. Similarly, another study (Ames et al., 2015) testing the effect of barley tortillas containing varying amounts and types of fibre on PPGR within 2 hours in 12 healthy volunteers found that there was a significant dose-dependent inverse effect of β-glucan (a soluble fibre, at 4.5 g, 7.8 g and 11.6 g per 50 g of available carbohydrate) on the PPGR while no significant difference in the PPGR between low and high insoluble fibre contents (7.4 g v. 19.6 g respectively). In addition, the microstructure properties of whole wheat flour doughs and their breads were reported significantly different from their refined version: the larger particle size and the widest spread of whole wheat bread make the starch more accessible to α-amylase activity, thus, the higher hydrolytic products of starch and high glycaemic response like white bread (Zafar et al., 2020).

Health professionals should be cautious when suggesting wholemeal bread over white bread to people particularly those with diabetes, who may consider wholemeal bread as a better option over white bread and thus consume it excessively without the expected health benefit of glycaemic control (Zafar et al., 2020). Meanwhile, bakery industries need to make innovative bread products that not only increase the dietary fibre content but also attenuate PPGR after consumption.

However, in contrast to the above results, a systematic review and meta-analysis (Marventano et al., 2017) found that the 14 studies testing the acute effects of wholegrain foods (mainly wholegrain rye bread) on PPGR showed significant reductions of the post-prandial iAUC (0–120 min) by −29.71 mmol min/L (95% CI: −43.57, −15.85 mmol min/L) compared to similar refined foods in healthy subjects. Like wholegrain wheat, wholegrain rye contains mainly insoluble dietary fibre (around 75%) (rye contains 15.2-20.9 g dietary fibre per 100 g among which 11.1-15.9 g are insoluble fibre and 3.7-4.5 g are soluble fibre (De Santis et al., 2018). The increased soluble fibre in wholegrain rye (around 25%) compared with wholegrain wheat (around 16%) (De Santis et al., 2018) might explain the difference in PPGR between wholegrain wheat and wholegrain rye bread consumption.

There is a trend to produce fibre-fortified innovative bread with the aim of increasing the dietary fibre intake of the population and reducing the glycaemic index (García et al., 2023). However, while the addition of wheat fibre or wheat bran can increase the total dietary fibre content in the bread, this does not necessarily deliver the health benefit of attenuating PPGR due to them being mainly insoluble fibre.

The current study used commercially available breads to represent commonly consumed staple foods and verified that the chosen wholemeal bread may not always provide the benefits claimed, especially regarding effects on blood glucose control. However, the result is only applicable to the breads chosen in the current study in an acute randomised crossover trial, and not applicable to other wholemeal breads or long-term consumption. In addition, this study had a small sample size (n=20) with diversity in gender and ethnicities in normal weight and overweight/obese groups We did not measure postprandial insulin; however, this would have been a useful tool for this study as research suggests this is the earliest biomarker for diagnosing pre-diabetes, T2DM and increased cardiovascular risk (DiNicolantonio et al., 2017). Further research should focus on the acute effect of different types and amounts of dietary fibre enriched bread on PPGR together with postprandial insulin response in a well-controlled population and/or long-term effect of bread consumption with different contents and types of dietary fibre in cohorts or randomised controlled trials.

## Data Availability

All data produced in the present study are available upon reasonable request to the authors.

## Acknowledgement

We sincerely thank Ms Susie Wilson, the Senior Technician at the School of Health and Life Sciences at Coventry University for her full training and support during the study period. We greatly appreciate the support from the UK Nutrition Society Summer Studentship scheme (Ref 71978).

## Authors’ contribution

HD, YX and AC designed the study. YX applied for the ethics approval. AC conducted the study under the supervision of YX. HD analysed the data and prepared the manuscript. HD had primary responsibility for the final content. All authors have read and approved the final manuscript.

The authors have no conflict of interest to report.

Data described in the manuscript will be made available upon request.

## References

Abdul-Ghani MA, Abdul-Ghani T, Ali N, Defronzo RA. One-hour plasma glucose concentration and the metabolic syndrome identify subjects at high risk for future type 2 diabetes. Diabetes Care. 2008;31(8):1650–1655. doi:10.2337/dc08-0225

Agrawall N, Agrawal MK, Kumari TN, & Kumar S. Correlation between Body Mass Index and Blood Glucose Levels in Jharkhand Population. Int. J. Contemp. Med. 2017; 4 (8): 1633–1636

Ames N, Blewett H, Storsley J, Thandapilly SJ, Zahradka P, Taylor C. A double-blind randomised controlled trial testing the effect of a barley product containing varying amounts and types of fibre on the postprandial glucose response of healthy volunteers. Br J Nutr. 2015;113(9):1373–1383. doi:10.1017/S0007114515000367

Belobrajdic DP, Regina A, Klingner B, et al. High-Amylose Wheat Lowers the Postprandial Glycaemic Response to Bread in Healthy Adults: A Randomized Controlled Crossover Trial. J Nutr. 2019;149(8):1335–1345. doi:10.1093/jn/nxz067

Chlup R, Seckar P, Zapletalová J, Langová K, Kudlová P, Chlupová K, Bartek J, & Jelenov, D. Automated computation of glycemic index for foodstuffs using continuous glucose monitoring. J Diabetes Sci Technol. 2008; 2(1): 67–75. 10.1177/193229680800200110

De Santis MA, Kosik O, Passmore D, Flagella Z, Shewry PR, Lovegrove A. Comparison of the dietary fibre composition of old and modern durum wheat (Triticum turgidum spp. durum) genotypes. Food Chem. 2018;244:304–310. doi:10.1016/j.foodchem.2017.09.143

DeFronzo RA, Ferrannini E, Groop L, Henry RR, Herman WH, Holst JJ, et al. Type 2 diabetes mellitus. Nat Rev Dis Prim. 2015;1:15019.

DiNicolantonio JJ, Bhutani J, OKeefe JH, Crofts C. Postprandial insulin assay as the earliest biomarker for diagnosing pre-diabetes, type 2 diabetes and increased cardiovascular risk. Open Heart. 2017;4(2):e000656. doi:10.1136/openhrt-2017-000656

García P, Bustamante A, Echeverría F, et al. A Feasible Approach to Developing Fiber-Enriched Bread Using Pomegranate Peel Powder: Assessing Its Nutritional Composition and Glycemic Index. Foods. 2023;12(14):2798. Published 2023 Jul 23. doi:10.3390/foods12142798

Giuntini EB, Sardá FAH, de Menezes EW. The Effects of Soluble Dietary Fibers on Glycemic Response: An Overview and Futures Perspectives. Foods. 2022;11(23):3934. Published 2022 Dec 6. doi:10.3390/foods11233934

Hannah B, Mallard S, Venn B. Glycemic Differences between White and Whole Grain Bread but No Differences in Glycemic Response between Sandwiches made with these Breads, Implications for Dietetic Advice. 2014; J Diabetes Metab. 5:456.

Johnson JD. On the causal relationships between hyperinsulinaemia, insulin resistance, obesity and dysglycaemia in type 2 diabetes. Diabetologia. 2021;64(10):2138–2146. doi:10.1007/s00125-021-05505-4

Juntunen KS, Laaksonen DE, Autio K, et al. Structural differences between rye and wheat breads but not total fiber content may explain the lower postprandial insulin response to rye bread. Am J Clin Nutr. 2003;78(5):957–964.

Klein S, Gastaldelli A, Yki-Järvinen H, Scherer PE. Why does obesity cause diabetes?. Cell Metab. 2022;34(1):11–20. doi:10.1016/j.cmet.2021.12.012

Leohr J, Kjellsson MC. Impact of Obesity on Postprandial Triglyceride Contribution to Glucose Homeostasis, Assessed with a Semimechanistic Model. Clin Pharmacol Ther. 2022;112(1):112–124. doi:10.1002/cpt.2604

Lockyer S, Spiro A. The role of bread in the UK diet: An update. Nutr Bull. 2020;45(2):133–164. 10.1111/nbu.12435

Marventano S, Vetrani C, Vitale M, Godos J, Riccardi G, Grosso G. Whole Grain Intake and Glycaemic Control in Healthy Subjects: A Systematic Review and Meta-Analysis of Randomized Controlled Trials. Nutrients. 2017;9(7):769. Published 2017 Jul 19. doi:10.3390/nu9070769

Moghaddam E, Vogt JA, Wolever TM. The effects of fat and protein on glycemic responses in nondiabetic humans vary with waist circumference, fasting plasma insulin, and dietary fiber intake [published correction appears in J Nutr. 2006 Dec;136(12):3084]. *J Nutr*. 2006;136(10):2506-2511. doi:10.1093/jn/136.10.2506

Monnier L, Lapinski H, Colette C. Contributions of fasting and postprandial plasma glucose increments to the overall diurnal hyperglycemia of type 2 diabetic patients: variations with increasing levels of HbA(1c). Diabetes Care. 2003;26(3):881–885. doi:10.2337/diacare.26.3.881

Musa-Veloso K, Poon T, Harkness LS, O’Shea M, Chu Y. The effects of whole-grain compared with refined wheat, rice, and rye on the postprandial blood glucose response: a systematic review and meta-analysis of randomized controlled trials. Am J Clin Nutr. 2018;108(4):759–774. doi:10.1093/ajcn/nqy112

NICE (2023) *Obesity: identification, assessment and management*. Online, available from https://www.nice.org.uk/guidance/cg189/ifp/chapter/obesity-and-being-overweight#:~:text=If%20you%20want%20to%20work,use%20an%20online%20BMI%20calculator.&text=Between%2018.5%20and%2024.9%20%E2%80%93%20healthy,Between%2030%20and%2039.9%20%E2%80%93%20obese (accessed on 03/08/2024)

Paiva AT, Gonçalves DR, Ferreira PS, et al. Postprandial effect of fresh and processed orange juice on the glucose metabolism, antioxidant activity and prospective food intake. J Funct Foods. 2019;52: 302–309. 10.1016/j.jff.2018.11.013

Perälä MM, Hätönen KA, Virtamo J, et al. Impact of overweight and glucose tolerance on postprandial responses to high- and low-glycaemic index meals. Br J Nutr. 2011;105(11):1627–1634. doi:10.1017/S0007114510005477

Raczkowska E, Bronkowska M. The Effect of the Body Mass Indexes of Young Healthy Individuals on the Glyacemic Indexes of Traditional and Modified Vegetarian Meals. Nutrients. 2019;11(10):2546. Published 2019 Oct 22. doi:10.3390/nu11102546

Shulman GI. Ectopic fat in insulin resistance, dyslipidemia, and cardiometabolic disease. N Engl J Med. 2014;371:1131–41.

Smith K, Taylor GS, Allerton DM, et al. The Postprandial Glycaemic and Hormonal Responses Following the Ingestion of a Novel, Ready-to-Drink Shot Containing a Low Dose of Whey Protein in Centrally Obese and Lean Adult Males: A Randomised Controlled Trial. Front Endocrinol (Lausanne). 2021;12:696977. Published 2021 Jun 18. doi:10.3389/fendo.2021.696977

van Vliet S, Koh HE, Patterson BW, et al. Obesity Is Associated With Increased Basal and Postprandial β-Cell Insulin Secretion Even in the Absence of Insulin Resistance. Diabetes. 2020;69(10):2112–2119. doi:10.2337/db20-0377

Vinoy S, Goletzke J, Rakhshandehroo M, et al. Health relevance of lowering postprandial glycaemia in the paediatric population through diet’: results from a multistakeholder workshop. Eur J Nutr. 2023;62(3):1093–1107. doi:10.1007/s00394-022-03047-y

Williams RA, Dring KJ, Cooper SB, Morris JG, Sunderland C, Nevill ME. Predictors of postprandial glycaemia, insulinaemia and insulin resistance in adolescents. Br J Nutr. 2021;125(10):1101–1110. doi:10.1017/S0007114520003505

Wunsch NG. Bread and bakery products in the UK - statistics and facts. 2022. Available online from https://www.statista.com/topics/7306/bread-and-bakery-products-in-the-uk/#dossierKeyfigures (accessed on 03/01/2025)

Zafar TA, Aldughpassi A, Al-Mussallam A, Al-Othman A. Microstructure of Whole Wheat versus White Flour and Wheat-Chickpea Flour Blends and Dough: Impact on the Glycemic Response of Pan Bread. Int J Food Sci. 2020; 2020:8834960. Published 2020 Oct 5. doi:10.1155/2020/8834960

